# Social determinants of health exacerbate disparities in COVID-19 illness severity and lasting symptom complaints

**DOI:** 10.1101/2021.07.16.21260638

**Authors:** Moriah E. Thomason, Cassandra L. Hendrix, Denise Werchan, Natalie H. Brito

**Author notes:** Corresponding Author: Moriah E. Thomason, NYU Grossman School of Medicine, Department of Child and Adolescent Psychiatry, One Park Ave, 8^th^ Floor, New York, NY 10016.

## Abstract

**BACKGROUND:** Increasing reports of long-term symptoms following COVID-19 infection, even among mild cases, necessitates systematic investigation into the prevalence and type of lasting illness. Notably, there is limited data regarding the influence of social determinants of health, like perceived discrimination and economic stress, which may exacerbate COVID-19 health risks. The primary goals of this study are to test the bearing of subjective experiences of discrimination, financial security, and quality of care on illness severity and lasting symptom complaints.

**METHODS:** 1,584 recovered COVID-19 patients that experienced mild to severe forms of the disease provided information about their illness, medical history, lasting symptoms, and psychosocial information. Prevalence data isolated differences in patients infected early versus late in the pandemic. Path analyses examined hypothesized associations between discrimination, illness severity, and lasting symptoms. *Post hoc* logistic regressions tested social determinants hypothesized to predict neurological, cognitive, or mood symptoms.

**RESULTS:** 70.6% of patients reported presence of one or more lasting symptoms after recovery. Neural systems were especially impacted, and 19.4% and 25.1% of patients reported mood or cognitive/memory complaints, respectively. Path models demonstrated that frequency and stress about experiences of discrimination predicted increased illness severity and increased lasting symptom count, even when adjusting for sociodemographic factors and mental/physical health comorbidities. Notably, this effect was specific to stress related to discrimination, and did not extend to general stress levels. Further, perceived but not objective socioeconomic status (SES) was associated with increased lasting symptom complaints after recovery. Finally, associations between discrimination and illness differed with individual perceptions about quality of medical care.

**CONCLUSIONS:** Lasting symptoms after recovery from COVID-19 are highly prevalent and neural systems are significantly impacted. Importantly, psychosocial factors (perceived discrimination and perceived SES) can exacerbate individual health risk. This study provides actionable directions for improved health outcomes by establishing that sociodemographic risk and medical care influence near and long-ranging health outcomes.

## Introduction

Reports of long-ranging symptoms following recovery from acute SARS-CoV-2 (COVID-19) infection have spurred widespread interest in the nature, prevalence and etiology of these lasting effects. A recent meta-analysis reports that up to 80% of patients developed one or more long-term symptom (14-110 days post-viral infection), with fatigue (58%), headache (44%), attention problems (27%), hair loss (25%) and dyspnea (24%) being the most frequent complaints. Recognition of medical complications that last weeks to months after initial recovery, so-called Long-COVID or COVID long-haulers, has led to rapid initiation of studies to rigorously study these effects. At the time of this writing, the US National Institutes of Health is launching a nationwide initiative, the Researching COVID to Enhance Recovery (RECOVER) study, that may soon involve ∼48 clinical sites, and the UK has launched the Post-Hospitalisation COVID-19 (PHOSP-COVID) study, involving ∼20 universities. Further, several independent groups are initiating much-needed deep-phenotyping studies in specialized populations, such as with children with pediatric inflammatory multisystem syndrome (MIS-C), to better understand special populations and/or better address indirect or complex pathways that contribute to disease outcomes.

Neurological properties of COVID-19 are gaining in importance and may be a vital aspect of research into long-COVID. It is unlikely that COVID-19 pathogens directly alter the brain; however, vascular walls and specific cells of the brain express angiotensin converting enzyme-2 (ACE2) and other docking receptors of SARS-Cov-2, suggesting vulnerability of the central nervous system (CNS) to the virus. However, expression of these receptors is relatively low and sensitivity of these receptors is variable.[1, 2] Moreover, COVID-19 virus is largely absent in patient CNS specimens obtained to date. Instead, data suggest that neurological complications associated with COVID-19 more likely arise from robust peripheral immune response to infection, secondary complications, and invasive therapies.[3, 4] For example, *ex vivo* studies confirm that COVID-19 infection is associated with elevated inflammatory markers, abnormal coagulation concentrations, increased cytokine expression, especially IL-6, IL-8 and TNF-alpha, endothelial dysfunction, hypercoagulable state, and imbalanced immune responses.[5, 6]

An important consideration in the study of lasting symptoms after recovery from COVID-19 is differential vulnerability. COVID-19 has had disproportionate impact on racial and ethnic minority groups, and it has been suggested that biomedical factors and social determinants of health underlie this difference.[7, 8] Empirical studies corroborate this mechanistic account, demonstrating that adjusting for sociodemographic factors and comorbidities in patients that reach medical care nullifies racial/ethnic differences.[9, 10] Thus, in the study of long-COVID, social and economic stress are key health determinants that must be addressed. In particular, it may be important to consider subjective experiences of discrimination and economic stress as risk factors that would elevate physical and neurological symptoms persisting or emerging after recovery from primary infection. For example, discrimination, in any form (e.g., disability, sexual orientation, physical appearance, race/ethnicity, religion) may act as a barrier to healthcare, further increasing risk of negative health outcomes due to underuse of mental health services, lower trust in healthcare systems, and delayed or avoided treatment.[11] Chronic stress resulting from perceived discrimination is also associated with allostatic load, [12, 13] which is defined as a dysregulation of the body’s physiological systems, including cardiovascular, neuroendocrine, metabolic, and immunologic systems that increases disease susceptibility and mortality. Despite the centrality of these factors, studies of lasting effects of COVID-19 infection have yet to address the role of prior health conditions, perceived equity, and stress, all of which are established critical determinants of health.

The present study was designed to address the prevalence, timing, and social determinants of lasting physical and neural symptoms in a large sample of patients that experienced mild to severe forms of COVID-19, months after recovery. We first provide a descriptive comparison between early and late COVID-19 cases, given the two-peak incidence of this pandemic. Primary analyses then test the hypothesis that frequency of and stress from discrimination contribute to COVID-19 illness severity and lingering symptoms in recovered patients, controlling for a variety of sociodemographic characteristics and mental and physical health morbidities. Secondary analyses address whether observed relationships are specific to stress associated with discrimination or reflect elevated stress more generally, and explore perceived quality of care as a potential buffer between predictors and outcomes. All analyses control for mental and physical health co-morbidities. *Post hoc* analyses test whether primary effects are predictive of a general syndrome of lasting effects or if there is evidence that neural domains are specifically affected. We examined these questions using first-person self-report data in a sample of 1,584 patients. All data have been made publicly available and curation/validation processes have been documented.[14]

## Methods

### Participants

The research protocol for this study was approved by the NYU Langone Institutional Review Board (IRB). A search of the NYU Langone Health record system in February 2021 identified 23,267 individuals ages 18 and older with COVID-19 diagnosis based on EPIC International Classification of Diseases (ICD-10) code U07.^*^ Of those on this list, individuals (1) with email contact, (2) not deceased, and (3) not designated as having previously opted out of research contact were eligible to participate. After application of these exclusions, 17,282 individuals were sent an email inviting them to participate in a 15-minute survey. Compensation was entry into a drawing for a $25 gift card. All surveys were completed between February 23, 2021 and April 4, 2021. Description of the survey measures and additional details on survey administration are described in ***Supplemental Material***. A total of 2,212 individual responses to the survey were received. 1,584 cases were retained after data validation measures were applied (see ***Supplemental Material***). For demographic characteristics of the final sample after quality assurance steps, see **Table S1**. For overview of sample illness timing and severity, see **Figure 1**.

**Figure 1.**
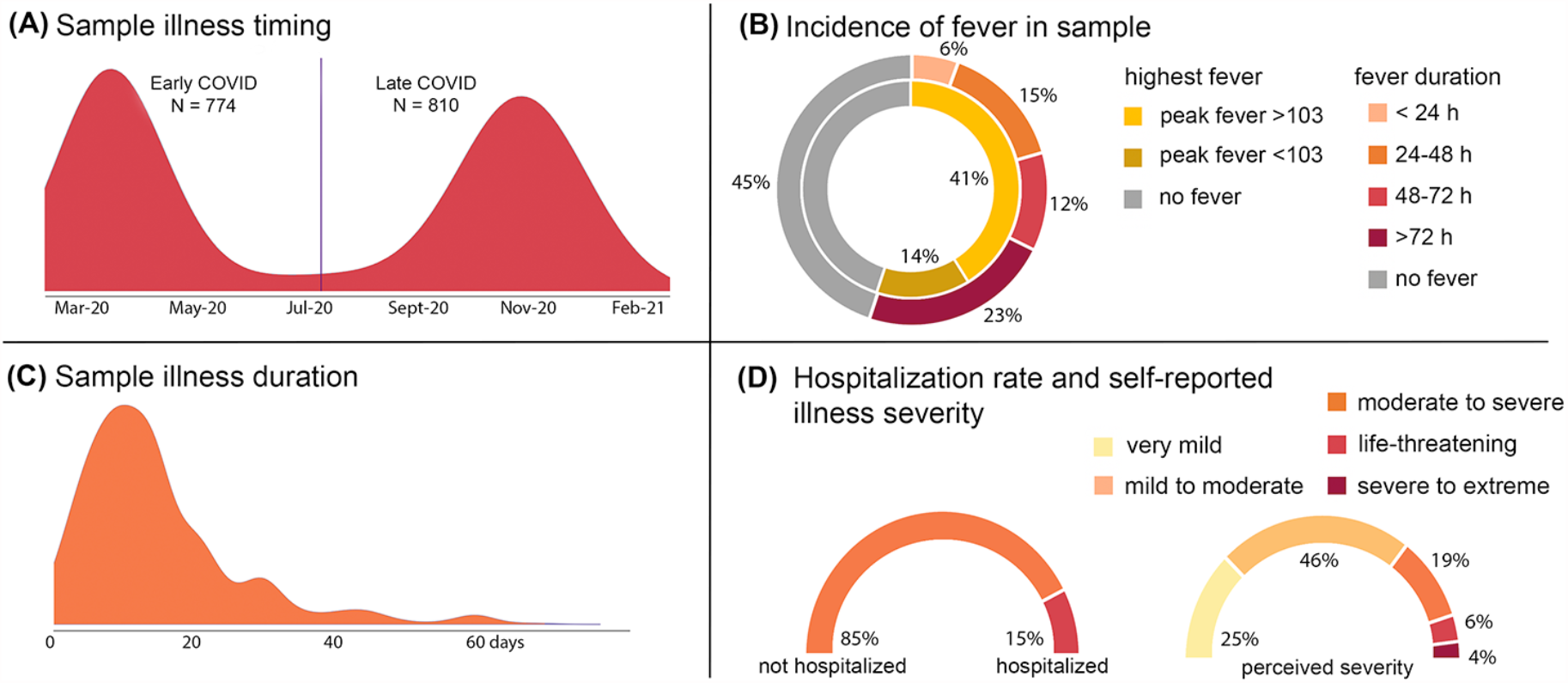
Overview of illness severity in current sample of N = 1,584 adults infected with COVID-19. Quality validated data were used to generate descriptive statistics that summarize illness timing (panel A); frequency and extent of fever (panel B); duration of illness (panel C); and both rate of hospitalization and self-reported illness severity in our adult sample. Bimodal distribution in panel A aligns well with observed incidence in New York City over this time period. The vertical line in panel A is the mean cut point used to analyze potential differences in early versus late infection groups.

### Statistical approach

A consideration is whether individuals that were ill during the first incidence peak of COVID-19 differ from individuals ill during the second peak in our sample. The peaks in NYC occurred on April 8, 2020 and on Jan 7, 2021.[15] The sample was mean split based on self-reported date of illness, resulting in split at July 24, 2020 for early versus late infection. Early and late cases were compared on sociodemographic, clinical, and psychosocial factors, using chi-squared tests and two-sample t-tests run with 5,000 bootstrapped samples.

Path analyses were used to test (1) direct associations between lifetime discrimination history, self-reported illness severity, and lasting symptom count, (2) indirect associations between discrimination and lasting symptoms, mediated through illness severity, and (3) moderation by stress from experiencing discrimination. Secondary analyses tested (1) specificity to discrimination stress, relative to experiencing increased stress in general, and (2) differences based on subjective perceptions of medical care (excellent versus non-excellent reported quality). See ***Supplemental Material*** for detailed descriptions of all measures.

Full information maximum likelihood (FIML) was used to avoid biased estimates associated with listwise deletion. Tests of statistical mediation were conducted using 5,000 bootstrap samples to generate bias-corrected confidence intervals. Race (white vs. non-white), objective SES risk score, perceived SES score, history of mood/anxiety disorder, history of diabetes or heart disease, COVID-illness life disruption, COVID-illness anxiety, and early versus late illness onset were controlled for in all analyses. All path analyses were conducted using Mplus v8.

## Results

### Prevalence and type of lasting symptom complaints

1,118 (70.6%) of participants reported presence of one or more lasting symptom after recovery from primary COVID-19 illness. Twenty-five percent of patients reported having cognitive or memory problems as a result of their COVID illness. Patients asked about kinds of cognitive complaints reported short-term memory (70%), attention (58%) and learning (22%) issues as the most frequently occurring. In addition, 19.4% of participants endorsed lasting mood symptoms following COVID-19 infection. The most common lasting mood complaints, in those that endorsed lasting mood complaints, were anxiety/nervousness (58%), depressed mood (19%) and irritation/short temper/agitation (15%). Frequencies of all observed lasting effects are provided in **Figure 2**.

**Figure 2.**
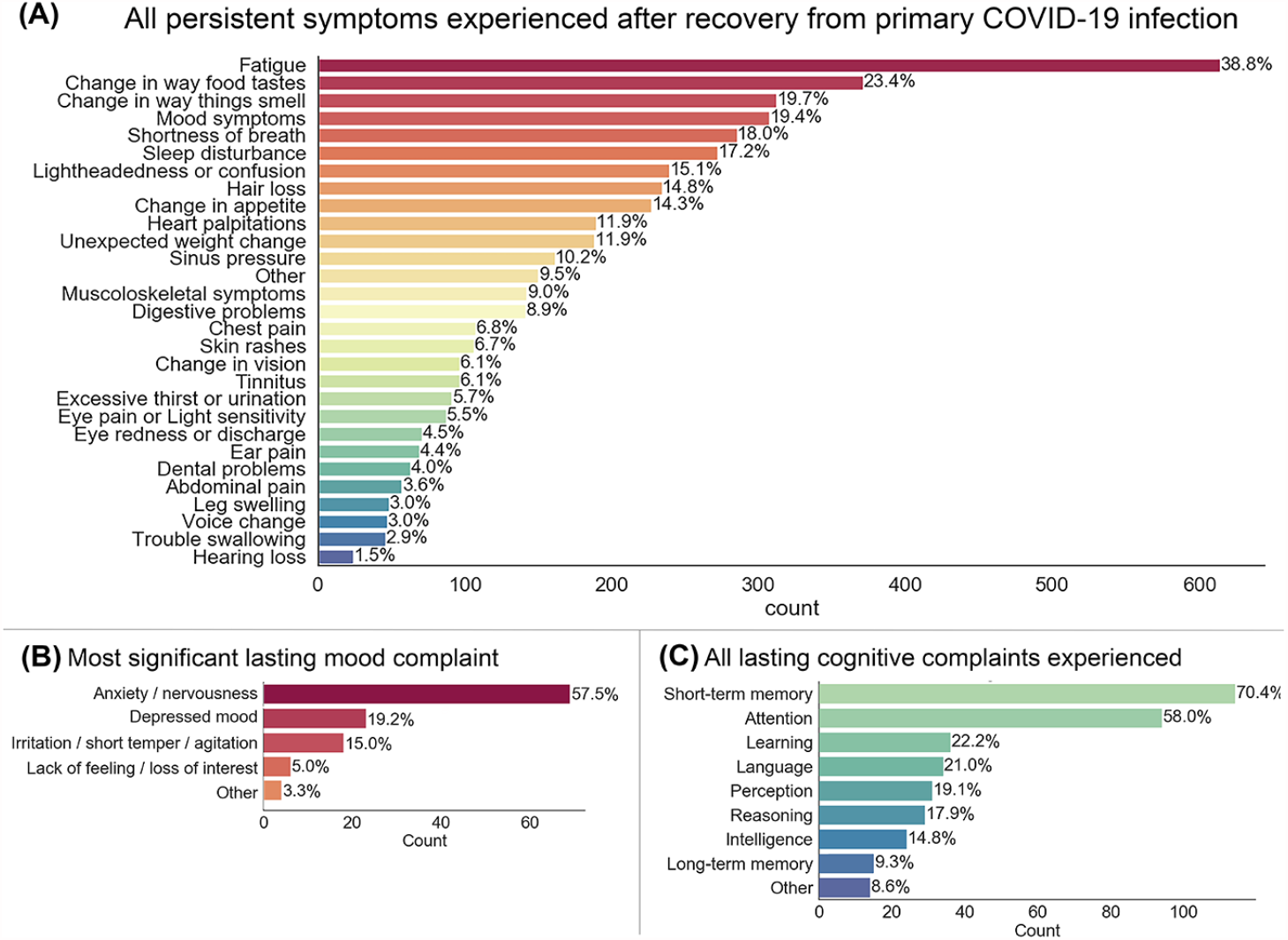
Prevalence of specific symptoms experienced by individuals reporting long-term sequelae following recovery from COVID-19. 70.6% of participants reported presence of one or more lasting symptom after recovery. The average number of lingering symptoms reported was 3.06 (SD = 3.73). Chief lingering symptom complaints in the sample were fatigue, change in the perception of taste and smell, and mood symptoms (panel A). Follow up questions in a subset of participants provide insight into the primary kinds of mood (panel B) and cognitive (panel C) complaints. The proportion of participants that reported mood or cognitive/memory complaints following illness were 19.4% and 25.1%, respectively.

### Early versus late timing of infection

Early versus late infection groups differed in illness severity (*p* = 5.3E-17), number of lasting symptoms (*p* = .002), number of lasting mood complaints after recovery (*p* = .00008), anxiety about illness (*p* = 9.2E-8) and ratings of COVID illness-related life disruption (*p* = .001). Direction of these effects was such that patients in the early infection group reported more severe illness, more lasting symptoms, and increased anxiety and life disruption. Full results of early versus late infection group comparisons are provided in ***Supplementary Material*, Table S2 and Figure S1**.

### Path model results

Results indicated that two important aspects of discrimination experiences – the frequency at which they occur and the stress associated with these experiences – interact to predict illness severity and lasting symptoms. In addition, the interactive effects of discrimination frequency and stress on lasting symptoms was partially mediated through illness severity (*β* = .02, CI [.01, .03]). In a model that did not include moderation by discrimination stress, the indirect association through illness severity was *not* present (*β* = −.01, CI [−.03, .01]). This indicates that illness severity explains the link between discrimination and lasting symptoms, and highlights the importance of considering individual differences in subjective stress from discrimination. Full path model results are reported in **Table S3** and illustrated in **Figure 3A**.

**Figure 3.**
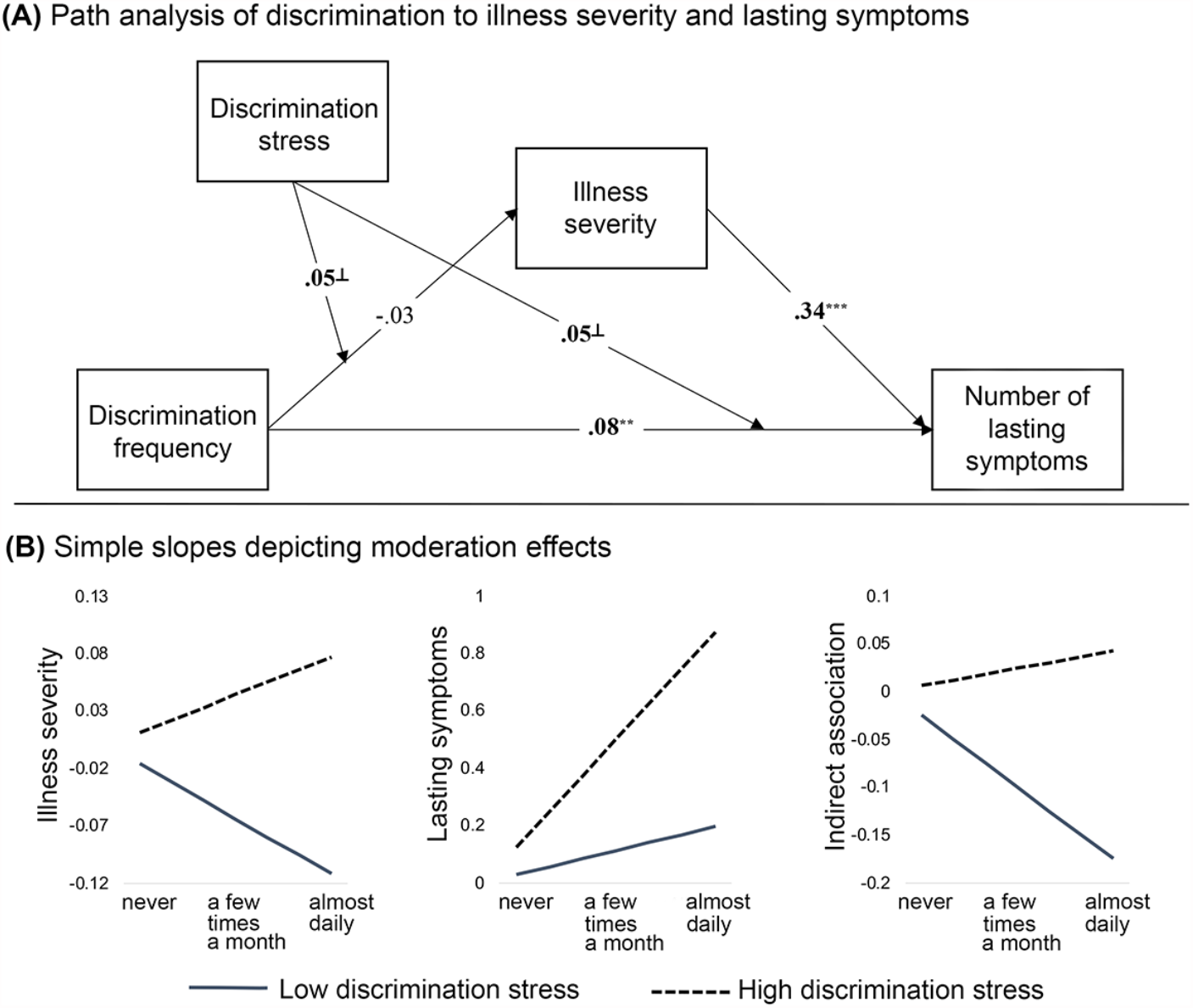
The observed path model and simple slopes depicting moderation effects. Observed associations between discrimination frequency, illness severity and number of lasting symptoms, with moderation by discrimination stress are represented in panel A. Standardized coefficients are shown. On all pathways, we controlled for race, cumulative SES risk score, perceived SES score, history of mood/anxiety disorder, history of diabetes/heart disease, COVID-illness life disruption, COVID-illness anxiety, and early versus late illness onset (i.e., peak 1 versus peak 2). A summary of observed moderation effects is provided in panel B, plotting model-estimated standardized simple slopes for all values of discrimination frequency. The x-axis for panel B is discrimination frequency. Discrimination stress moderates the direct effects of discrimination frequency on illness severity and lasting symptoms (left and middle plot). Discrimination stress also moderates the indirect effect of discrimination frequency on lasting symptoms through differential impacts on illness severity (right plot). ^⊥^ *p* < .10, ^**^ *p <* .01, ^***^ *p <* .001.

Simple slopes for individuals 1 SD +/− the mean for discrimination stress provide a visual representation of the direction of observed effects. Slopes indicate that increased frequency of discrimination was a stronger predictor of both increased illness severity and increased lasting symptoms for individuals reporting higher levels of stress from discrimination, see **Figure 3B**. Further, there was a positive mediation effect of increased discrimination on increased lasting symptoms through greater illness severity for individuals reporting higher stress from discrimination (mean +1 SD: *β* = .01, CI [−.01, .02]), but not for individuals reporting lower stress from discrimination (mean −1 SD: *β* = −.03, CI [−.05, −.003]); see **Figure 3B**. These findings suggest that both the frequency of and stress associated with chronic discrimination contribute to disparities in COVID-19 health outcomes.

It is possible, however, that observed effects may not be specific to stress associated with chronic experiences of discrimination, but may instead be driven by poorer outcomes associated with increased stress levels more generally. As an analytical control, the same path analysis was tested using current stress levels as a moderator, instead of discrimination stress, which was included as an additional covariate. Results indicated that current stress moderated the impact of discrimination frequency on lasting symptoms (**Figure S2**). However, current stress did not moderate the direct effect of discrimination frequency on illness severity, nor the indirect association between discrimination frequency and lasting symptoms through illness severity. These findings suggest that chronic stress associated with frequent experiences of discrimination has a unique contribution to predicting disparities in health outcomes.

### Associations between discrimination and illness differ with individual perceptions of clinical care quality

A path analysis examined whether perceived quality of care influenced the observed conditional associations, as depicted in **Figure 3**. Detailed coverage of this analysis is provided as ***Supplementary Material***. In brief, participants were divided into high and low quality of care groups, based on self-ratings of care as excellent (n = 727) or less than excellent (n = 740). The majority of paths remained significant when looking within the low quality of care group; however, many of these paths were no longer significant when looking only within the high quality of care group (**Figure S3**). This suggests that high quality of perceived clinical care may impact links between chronic discrimination and illness severity.

### Lasting neurological syndrome

In final, exploratory analyses, we used logistic regression to address whether higher perceived discrimination was predictive of a general syndrome of lasting effects, or if there is evidence that neural domains are specifically affected. In these exploratory analyses, we also addressed whether perceived SES, COVID-life disruption, or illness-related anxiety predicted occurrence of lasting neural symptoms. We observed a significant positive effect of both discrimination frequency, **Figure 4A**, and COVID-life disruption, *B* = .16, *SE* = .12, *β* = .19, *p* = .01 (not pictured), on number of lasting neurological symptoms. Discrimination frequency was not predictive of lasting cognitive symptoms, *B* = .05, *SE* = .06, *β* = .06, *p* = .47; however, perceived SES, **Figure 4C**, and COVID-life disruption, *B* = .32, *SE* = .08, *β* = .40, *p* < .001 (not pictured), both had a significant effect on presence of lasting cognitive symptoms. Finally, a trend was observed in the association between lasting mood symptoms and discrimination frequency, **Figure 4B**, and significant positive effects for illness anxiety, *B* = .34, *SE* = .09, *β* = .46, *p* < .001 and COVID-life disruption, *B* = .27, *SE* = .09, *β* = .33, *p* = .01 (not pictured), with lasting mood symptoms.

**Figure 4.**
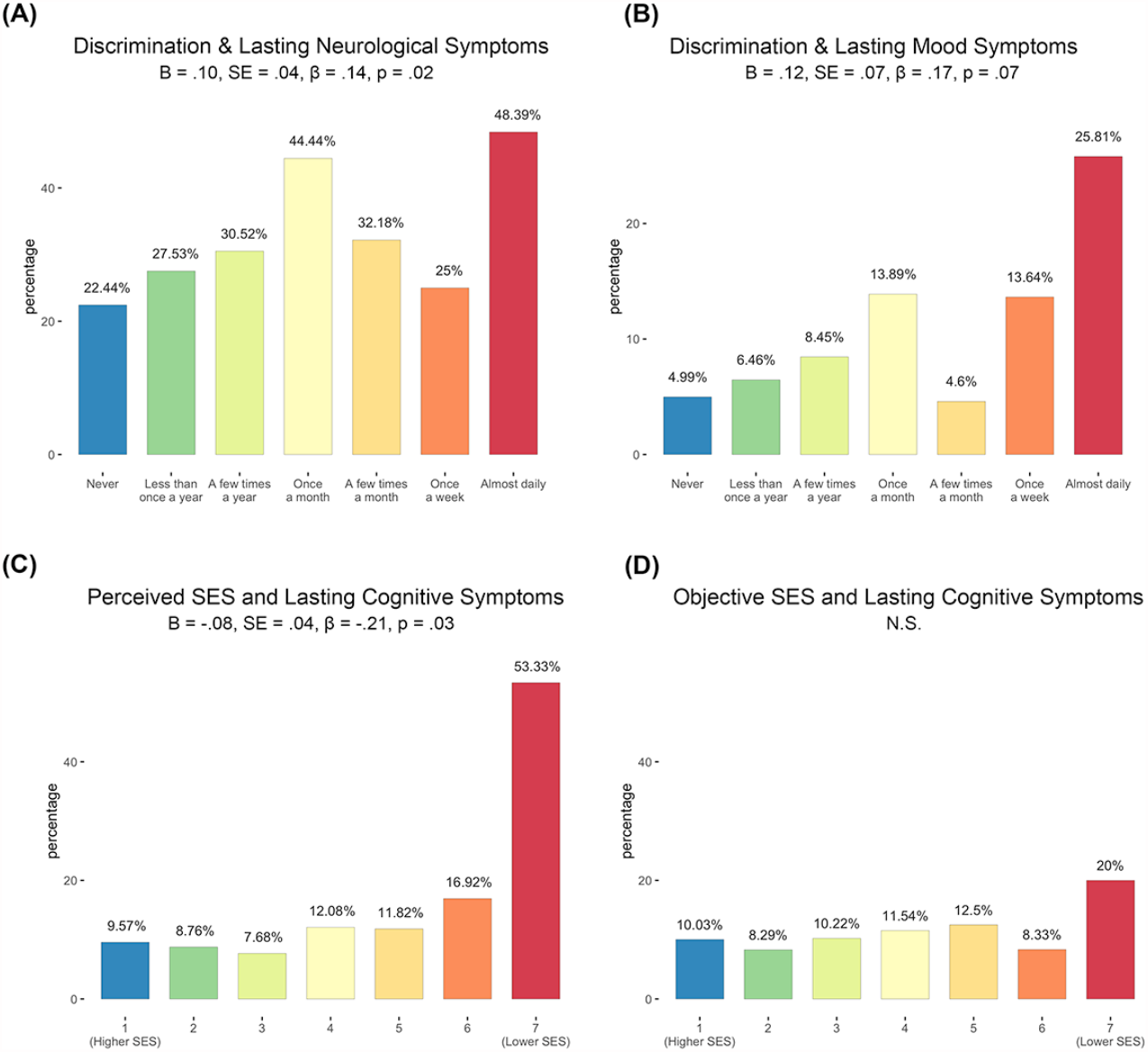
Perceived discrimination and perceived SES predict increased neurological and cognitive symptoms after recovery from COVID-19. *Post hoc* logistic regressions provide evidence that individuals reporting greater discrimination frequency had a significantly greater likelihood of reporting lasting neurological symptoms, *p* = .02 (panel A). Further, data demonstrate that perceived (panel C) but not objective (panel D) SES predicts lasting cognitive complaints after recovery from COVID-19 illness. There was a trend in the relationship between increased discrimination frequency and lasting mood symptoms, *p* = .07 (panel B). All analyses controlled for non-white race, cumulative SES risk score, perceived SES score, history of diabetes/heart disease, COVID-illness life disruption, COVID-illness anxiety, and early versus late illness onset (i.e., peak 1 versus peak 2).

## Discussion

Primary conclusions from this study are that: (1) lasting symptoms are common, with 70.6% of patients in this study reporting one or more symptoms, (2) specific psychosocial factors (perceived discrimination and perceived SES) place select individuals at greater health risk, (3) neural systems are significantly impacted, (4) patient perceptions regarding quality of medical care can be important in interpreting these relationships, and (5) illness early in the pandemic is associated with more severe illness and more frequent lasting complaints. These findings were obtained in a large sample of patients ranging in age from 18 to 96 years old, with varied health backgrounds, and with illness severity ranging from mild to severe. While not central to the present study, we also find that age and sex of patients also relate to number of lasting symptoms after recovery (see ***Supplemental Material***).

A major focus of this project was to evaluate individual determinants of long-COVID, with specific attention on experiences of discrimination as a predictor of health outcomes. Data presented suggest that chronic discrimination is a significant predictor of lasting COVID-19 sequalae through both direct and indirect pathways, in models that account for mental and physical health comorbidities and sociodemographic factors. The addition of discrimination stress, but not current stress, to path models affects the association between chronic discrimination and illness severity, highlighting specificity of observed effects. Thus, it is not a general syndrome of increased psychosocial burden; instead, it appears that frequent experiences of general discrimination place individuals at greater risk for becoming more ill when infected, and at greater risk for experiencing increased lasting health complaints after recovery.

A notable takeaway from the present study is the importance of subjective versus objective perspectives. This is illustrated well in the finding that perceived SES was a more robust predictor of long-term outcomes than actual SES, both of which were composite factors with high internal validity. Another notable observation was that associations between experiences of discrimination and illness differed based on the patient’s perceived quality of medical care. This is an encouraging avenue for promoting health in individuals at enhanced risk.

Overall, this study highlights the urgency for research to rigorously address long-term physical and neurological outcomes of COVID-19, as a large proportion of our global population has now been infected. Knowledge about the primary compromised domains informs our approach to treating afflicted individuals. Future studies would benefit from collecting information about patient perceptions and experiences, as these are clearly significant drivers of health outcomes. Knowledge about predictors and prevalence of lasting illness sequelae makes it possible to make informed economic and policy decisions about research and treatment.

## Data Availability

The primary dataset is accessible via the Open Science Framework (OSF) open access platform. Included in the release are: (a)raw data (.csv), (b)the NCIPR questionnaire (.pdf), (c)REDCap instrument files (.zip), and (d)the variable definition file (.csv). See https://osf.io/82rkj/

## Acknowledgements

This project was supported by National Institutes of Health awards DA050287, MH126468, and MH122447 to MET and NHB. The authors thank Autumn Austin, Carly Lenniger, Tessa Vatalaro, Integra Feliciano, Nicki Jariwala, Amyn Majbri, Harini Srinivasan, and Sarwat Siddiqui for contributions to recruitment and data collection. The authors thank participants who generously shared their time and who expressed interest in helping researchers and clinicians better understand the varied experiences and circumstances surrounding COVID-19 illness and recovery.

## Author contributions

MT designed the study and contributed materials. DW and CH analyzed the data. MT, DW, CH, and NB conceptualized and wrote the paper.

## References

1. Chen, R., et al., The Spatial and Cell-Type Distribution of SARS-CoV-2 Receptor ACE2 in the Human and Mouse Brains. Front Neurol, 2020. 11: p. 573095.

2. Brewster, L.M. and Y.K. Seedat, Why do hypertensive patients of African ancestry respond better to calcium blockers and diuretics than to ACE inhibitors and β-adrenergic blockersã A systematic review. BMC Med, 2013. 11: p. 141.

3. Boldrini, M., P.D. Canoll, and R.S. Klein, How COVID-19 Affects the Brain. JAMA Psychiatry, 2021.

4. Iadecola, C., J. Anrather, and H. Kamel, Effects of COVID-19 on the Nervous System. Cell, 2020. 183(1): p. 16-27.e1.

5. Mukerji, S.S. and I.H. Solomon, What can we learn from brain autopsies in COVID-19ã Neurosci Lett, 2021. 742: p. 135528.

6. Bryce, C., et al., Pathophysiology of SARS-CoV-2: targeting of endothelial cells renders a complex disease with thrombotic microangiopathy and aberrant immune response. The Mount Sinai COVID-19 autopsy experience. medRxiv, 2020: p. 2020.05.18.20099960.

7. Ajilore, O. and A.D. Thames, The fire this time: The stress of racism, inflammation and COVID-19. Brain Behav Immun, 2020. 88: p. 66–67.

8. Tai, D.B.G., et al., The Disproportionate Impact of COVID-19 on Racial and Ethnic Minorities in the United States. Clin Infect Dis, 2021. 72(4): p. 703–706.

9. Yehia, B.R., et al., Association of Race With Mortality Among Patients Hospitalized With Coronavirus Disease 2019 (COVID-19) at 92 US Hospitals. JAMA Netw Open, 2020. 3(8): p. e2018039.

10. Ogedegbe, G., et al., Assessment of Racial/Ethnic Disparities in Hospitalization and Mortality in Patients With COVID-19 in New York City. JAMA Netw Open, 2020. 3(12): p. e2026881.

11. Nong, P., et al., Patient-Reported Experiences of Discrimination in the US Health Care System. JAMA Netw Open, 2020. 3(12): p. e2029650.

12. Van Dyke, M.E., et al., Pervasive Discrimination and Allostatic Load in African American and White Adults. Psychosom Med, 2020. 82(3): p. 316–323.

13. Miller, H.N., et al., The impact of discrimination on allostatic load in adults: An integrative review of literature. J Psychosom Res, 2021. 146: p. 110434.

14. Thomason, M., D. Werchan, and C. Hendrix, COVID-19 patient accounts of illness severity, treatments and lasting symptoms. Scientific Data, 2021.

15. https://www1.nyc.gov/site/doh/covid/covid-19-data-trends.page.

