## supplemental material for "Social determinants of health exacerbate disparities in COVID-19 illness severity and lasting symptom complaints"

### *Table of Contents*

**I.** Data validation and exclusion criteria
**II.** Participant overview (**Table S1**)
**III.** Survey measures
    Overview of Novel Coronavirus Illness Patient Report (NCIPR) survey
    Derivation of summary measures
        COVID-19 illness severity
        SES risk score (objective variables)
        Perceived SES risk
        Perceived Discrimination (**Table S2**)
**IV.** Assessment of lasting symptoms
    Overall prevalence
    Prevalence of lasting mood and cognitive complaints
**V.** Temporal analysis of early and late COVID-19 cases (**Table S3** and **Figure S1**)
**VI.** Path model results (**Table S4**)
**VII.** Stress as an analytical control (**Figure S2**)
**VIII.** Quality of care as a protective factor (**Figure S3**)
**IX.** Most troubling symptoms experienced during COVID-19 illness (**Figure S4**)
**X.** Age and sex of patient relate to number of lasting symptoms after recovery from COVID-19 illness (**Figure S5**)

**Data validation and exclusion criteria**

Data assurance and quality checking were performed using R version 4.0.2. Quality of patient responses were evaluated by isolating implausible and/or inconsistent responses. For example, a height feet value greater than 7, or a height inches value greater than 12 were considered implausible, and conflicting self-reported 'date of birth' (DOB) and 'current age' were considered inconsistent. Respondents with implausible or inconsistent responses were excluded (N=72). Exclusions were also made on the basis of self-reporting not being ill with COVID-19 (N=65), incomplete survey data (N=483), and date of illness provided falling outside of February 2020 to March 2021. All data presented herein are based on the resulting sample of 1,584 recovered patients described in **Table S1,** below. Variation in illness severity in the sample is depicted in **Figure 1, main text**.

**Participant overview**

**Table S1.** Patient demographics after data validation

| | N =1,584<br>M(SD) or N(%) |
|---|---|
| *Age in years* | 44.64 (14.99) |
| *Education* | |
| Less than 10th grade | 5 (0.3%) |
| 10th-12th grade | 8 (0.5% |
| High school degree/GED | 77(5.0%) |
| Trade school/apprenticeship | 30 (1.9%) |
| Partial college | 141 (9.1%) |
| 2-year college degree | 117 (7.6%) |
| 4-year college degree | 548 (35.5%) |
| Graduate degree | 618 (40.0%) |
| *Gender* | |
| Female | 1115 (70.4%) |
| Male | 465 (29.4%) |
| Nonbinary | 3 (0.2%) |
| *Partnered/married* | 861 (54.5%) |
| *Race/Ethnicity* | |
| White | 994 (66.4%) |
| Hispanic/Latin | 164 (10.9%) |
| Black/African American | 121(8.1%) |
| Asian | 120 (8.0%) |
| Mixed | 76 (5.1%) |
| Other | 19 (1.2%) |
| Native American/Native Alaskan | 4 (<0.1%) |
| Native Hawaiian/Pacific Islander | 0 (0%) |
| *Previous mental health treatment* | 396 (25.0%) |
| *Pre-existing medical conditions* | 730 (46.1%) |
| Mood or anxiety disorder | 288 (18.2%) |
| Heart disease or hypertension | 244 (15.4%) |
| Respiratory problems | 228 (14.4% |
| Diabetes | 119 (7.5%) |
| Cancer | 118 (7.4%) |
| Lung disease | 34 (2.1%) |
| Liver disease | 24 (1.5%) |
| *Number of lasting symptoms* | 3.06 (3.73) |

**COVID-19 Illness severity**

The Novel Coronavirus Illness Patient Report (NCIPR) Survey was developed in November 2020 and published in the U.S. National Library of Medicine (NLM) Disaster Management Resources (id:24224) as well as the Open Science Framework (https://osf.io/82rkj/) in early March 2021. NCIPR was created as a means of collecting first-person accounts of COVID-19 illness and treatment, proximal to the experience of being ill. NCIPR includes questions addressing symptoms, medical complications, home and hospital treatments, lasting effects, anxiety about illness, employment impacts, quarantine behaviors, vaccine-related behaviors and effects, and illness of other family/household members. Additional questions address pandemic impacts (relationship, social, stress, sleep), health history, and coping strategies. A summary of all domains measured is described with the recent release of the dataset.[1] For the present study, NCIPR was used to derive a summary measure of COVID-19 illness severity. Illness severity was computed by standardizing and averaging the following relevant variables from the NCIPR instrument: fever severity, illness length, self-reported illness severity, medical complications, and COVID-related hospitalization. Confirmatory factor analysis for this composite NCIPR illness severity measure yielded $RMSEA = 0.085$, $CFI = .971$, $\chi 2 = 2003.33$, $p < 0.001$.

**Assessment of lasting symptoms, life disruption and illness anxiety**

The NCIPR survey includes questions about a broad array of lasting symptoms (n = 28), covering multiple functional systems, and also questions specific to cognitive/memory and mood complaints following infection. Magnitude of lasting symptoms was determined by summing the number of endorsed lasting complaints, and for *post hoc* analyses, lasting complaints were subdivided in to general complaints and central nervous system (CNS) specific complaints. The latter included fatigue, cognitive symptoms, mood symptoms, change in the way things taste, change in the way things smell, and sleep disturbance. In addition, individual NCIPR questions addressed COVID-illness life disruption and COVID-illness anxiety using a 6-point Likert scale ranging from none (0) to extreme (5).

**Discrimination**

Subjective experiences of discrimination were assessed using questions from the Perceived Discrimination Scale by Williams and colleagues.[2] Participants answered the following multiple choice items: (1) In your day-to-day life, have you experienced discrimination?; (2) What do you think was the reason(s) for this/these experience(s)? Pick as many as apply (check all that apply); and (3) Over your entire lifetime, how stressful have experiences of unfair treatment or discrimination usually been for you? Heading and item response choices are provided in **Table S2**.

**Table S2.** Questions assessing perceived discrimination frequency, type, and associated stress

**Instructions**

These are some questions about discrimination that you may or may not experience in your day-to-day life. By discrimination, we mean being treated unfairly because of your race, ethnicity, income level, social class, sex, gender, age, sexual orientation, physical appearance, or religion.

**Q1. In your day-to-day life, have you experienced discrimination?**

- Never
- Less than once a year
- A few times a year
- A few times a month
- Once a month
- At least once a week
- Almost every day
- I decline to answer

**Q2. What do you think was the reason(s) for this/these experience(s)? Pick as many as apply *(check all that apply)***

- Your ancestry or national origin
- Your gender
- Your race
- Your age
- Your height
- Your weight
- Some other aspect of your physical appearance
- Your sexual orientation
- Income Level/Social Class
- Disability
- Pregnancy
- Religion
- Some other reason *(if other, please specify:)*
- None apply to me
- I decline to answer

**Q3. Over your entire lifetime, how stressful have experiences of unfair treatment or discrimination usually been for you? *(check one)***

- Not at all stressful
- A little stressful
- Somewhat stressful
- Extremely stressful
- I decline to answer

Source: Williams, D.R., et al., Racial Differences in Physical and Mental Health: Socio-economic Status, Stress and Discrimination. J Health Psychol, 1997. 2(3): p. 335-51.

### Socioeconomic status

A measure of cumulative *objective* socioeconomic status (SES) was generated by standardizing and summing the following demographic variables: household income-to-needs ratio (i.e., income relative to household size), education level, stability of housing, and receipt of public assistance. Next, a measure of *perceived* SES was computed by standardizing and summing financial satisfaction, financial worries, perceived financial stability, and the MacArthur ladder of perceived social standing. Confirmatory factor analyses were used to verify fit of SES composite variables, which indicated excellent fit for both variables (objective SES: *RMSEA* = 0.029, *CFI* = .985, $\chi 2$ = 177.53, $p < 0.001$; perceived SES: *RMSEA* = 0.0, $\chi 2$ = 632.03, $p < 0.001$). Demographic questions and response options used to derive these measures are available at: https://osf.io/82rkj, via the NCIPR Demographic Survey.

### Temporal analysis of early and late COVID-19 cases

**Table S3.** Sociodemographic and illness characteristics in patients infected early or late in the pandemic

| | Early COVID (n=774) M(SD) or N(%) | Late COVID (n=810) M(SD) or % | Statistical results |
|---|---|---|---|
| *Sociodemographic* | | | |
| Age in years | 44.42 (14.25) | 44.85 (15.66) | $t$(1543)=-0.56, $p$=0.58, *95CI*[-1.98, 0.99] |
| Education | 6.90 (1.41) | 6.78 (1.44) | $t$(1542)=1.70, $p$=0.09, *95CI*[-0.01, 0.27] |
| Female gender | 552 (71.4%) | 563 (69.5%) | $X^2$(3,1583)=0.92, p=0.63 |
| Partnered/married | 254 (54.4%) | 440 (54.3%) | $X^2$(1,1584)=3.00, p=0.70 |
| Non-White race | 267 (34.5%) | 237 (29.3%) | $X^2$(1,1584)=5.00, p=0.03 |
| Composite SES risk score | 0.01 (2.36) | -0.01 (2.26) | $t$(1582)=0.08, $p$=0.93, *95CI*[-0.22, 0.23] |
| *Clinical characteristics* | | | |
| Previous mental health treatment | 190 (24.5%) | 206 (25.4%) | $X^2$(1,1584)=0.17, p=0.69 |
| Any pre-existing medical conditions | 356 (46.0%) | 374 (46.2%) | $X^2$(1,1584)=0.01, p=0.94 |
| [a] **Composite illness severity** | **0.13 (0.82)** | **-0.17 (0.58)** | **$t$(1384)=8.49, $p$=5.3E-17, *95CI*[0.23, 0.37]** |
| [a] **Lasting symptoms after recovery *(range 0-29)*** | **3.33 (4.00)** | **2.76 (3.42)** | **$t$(1522)=3.10, $p$=0.002, *95CI*[0.22, 0.96]** |
| Lasting neurological changes after recovery *(range 0-6)* | 0.44 (0.74) | 0.43 (0.73) | $t$(1582)=0.29, p=0.77, *95CI*[-0.06, 0.08] |
| Lasting cognitive/memory problems after recovery | 85 (11.0%) | 77 (9.5%) | $X^2$(3,1584)=5.23, p=0.16 |
| [a] **Lasting mood complaints after recovery** | **181 (23.4%)** | **126 (15.6%)** | **$X^2$(1,1584)=15.23, p=0.00008** |
| Lasting symptoms ongoing | 348 (45.0%) | 367 (45.3%) | $X^2$(1,1584)=0.02, p=0.89 |
| Were you satisfied with the medical care you received? *(1=very satisfied, 5=very dissatisfied)* | 2.01 (1.12) | 1.87 (1.07) | $t$(1465)=2.35, $p$=0.02, *95CI*[0.02, 0.25] |
| *Psychosocial environment* | | | |
| Discrimination frequency | 2.66 (1.41) | 2.47 (1.41) | $t$(1398)=2.57, $p$=0.01, *95CI*[0.05, 0.35] |
| Discrimination stress | 2.12 (0.94) | 2.03 (0.97) | $t$(1380)=1.79, p=0.07, *95CI*[-0.01, 0.19] |
| Perceived SES | -0.08 (2.43) | 0.07 (2.39) | $t$(1582)=-1.23, $p$=0.22, *95CI*[-0.38, 0.10] |
| [a] **How anxious were you about being ill? *(0=no anxiety, 5=extreme anxiety)*** | **2.18 (1.39)** | **1.82 (1.32)** | **$t$(1582)=5.37, p= 9.2E-8, *95CI*[0.23, 0.50]** |
| Please rate current stress level *(1 = nothing, 7 = extreme)* | 4.07 (1.49) | 3.93 (1.52) | $t$(1491)=1.77, p=0.08, *95CI*[-0.02, 0.29] |
| [a] **How much did your COVID illness disrupt your life? *(0=no disruption, 5=extreme disruption)*** | **2.52 (1.29)** | **2.31 (1.20)** | **$t$(1560)=3.35, p=0.001, *95CI*[0.09, 0.34]** |

[a] *< .05 after application of Holm–Bonferroni correction for multiple comparisons*

Differences in early versus late infection timing groups are reported in **Table S3.** Groups did not differ in sociodemographic variables, including age, education, gender, marital status, race, or objective SES. In addition, groups did not differ in prevalence of prior mental health treatment, pre-existing medical conditions, lasting neurological and/or cognitive symptoms after recovery, presence of ongoing lasting symptoms, perceptions about quality of care, perceived discrimination, perceived SES, or current stress. However, significant differences in early versus late infection groups were observed for composite illness severity ($p$ = 5.3E-17), number of lasting symptoms ($p$ = .002), and number of lasting mood complaints after recovery ($p$ = .00008). Direction of these effects was such that patients in the early infection group reported more severe illness and more long-COVID symptoms. Early versus late groups were also significantly different in anxiety about illness ($p$ = 9.2E-8) and ratings of COVID illness-related life disruption ($p$ = .001), where the early infection group reported increased concerns anxiety and life disruption. Analyses controlled for multiple comparisons (see **Table S3**). Select outcomes are plotted for early and late infection groups in **Figure S1**, below.

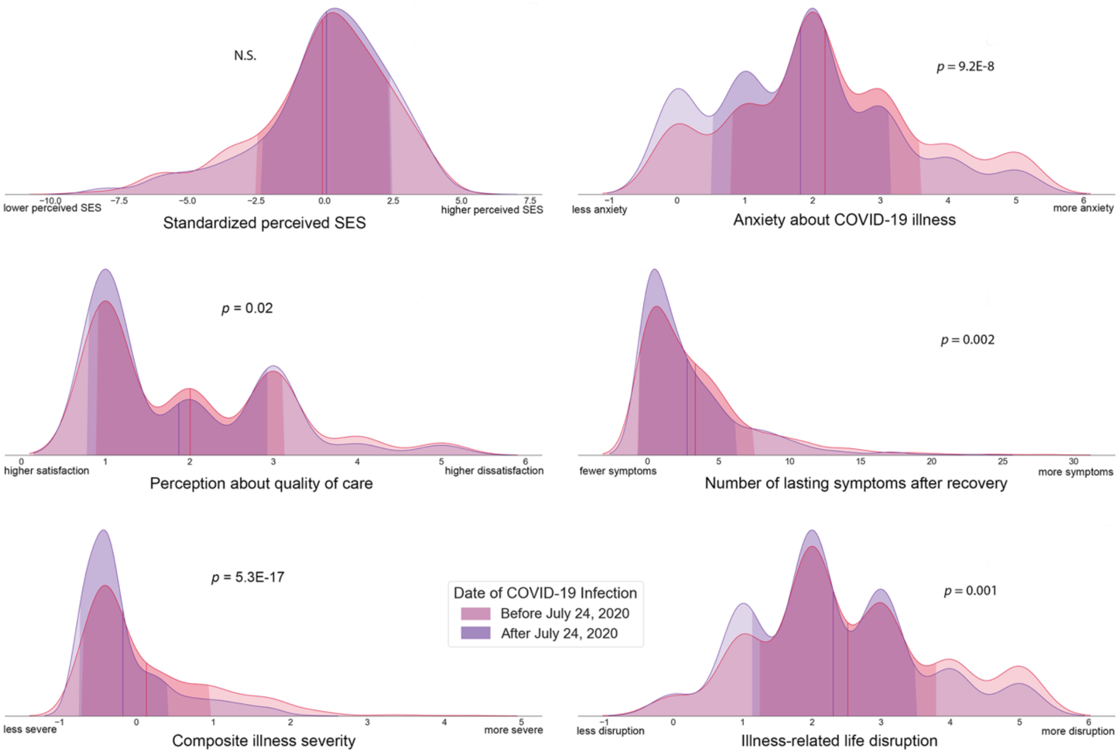


**Figure S1. Differences in patients infected early versus late in the COVID-19 pandemic.** Comparisons between patients infected with COVID-19 early versus late in the pandemic yield mixed results. While groups do not differ greatly in demographics, there are a few pronounced differences in clinical and psychosocial factors, most notably self-reported illness severity and anxiety about COVID-19 illness. Vertical red and purple lines on each distribution plot represent group means for early versus late infection, respectively. Standard deviations for each group are indicated by the darker shading on each plot.

### Path model results

Full results from the observed path model are reported in **Table S4.**

**Table S4.** Path analysis results for model 1 (including moderation by discrimination stress)

| Direct paths | *β (SE)* | *p* |
|---|---|---|
| *Number of Lasting Symptoms* | | |
| Discrimination Frequency | .08 (.03) | 0.01 |
| Discrimination Stress | .07 (.03) | 0.01 |
| Discrimination Frequency X Discrimination Stress | .05 (.03) | 0.09 |
| Illness Severity | .34 (.03) | < .001 |
| Non-White Race [a] | -.03 (.02) | 0.17 |
| Objective SES Score | .06 (.03) | 0.03 |
| Perceived SES Score | -.05 (.03) | 0.05 |
| Mood/Anxiety Disorder History | .05 (.03) | 0.06 |
| Diabetes/Heart Disease History | .01 (.03) | 0.8 |
| COVID-Illness Life Disruption | .13 (.03) | < .001 |
| COVID-Illness Anxiety | .09 (.03) | < .001 |
| Early vs Late Illness Onset | .02 (.02) | 0.26 |
| *Illness Severity* | | |
| Discrimination Frequency | -.03 (.03) | 0.3 |
| Discrimination Stress | -.03 (.03) | 0.34 |
| Discrimination Frequency X Discrimination Stress | .05 (.10) | 0.06 |
| Non-White Race [a] | .03 (.02) | 0.28 |
| Objective SES Score | .10 (.03) | < .001 |
| Perceived SES Score | .03 (.03) | 0.33 |
| Mood/Anxiety Disorder History | -.03 (.02) | 0.15 |
| Diabetes/Heart Disease History | .14 (.03) | < .001 |
| COVID-Illness Life Disruption | .45 (.03) | < .001 |
| COVID-Illness Anxiety | -.01 (.03) | 0.85 |
| Early vs Late Illness Onset | -.17 (.02) | < .001 |
| **Indirect paths** | **Estimate** | **95% CI** |
| Discrimination Frequency → Illness Severity → Lasting Symptoms | -0.01 | [-.03, .01] |
| Discrimination Frequency X Stress → Illness Severity → Lasting Symptoms | 0.02 | [.003, .03] |

[a] *Non-white race was dummy-coded (0 = white, 1 = non-white)*

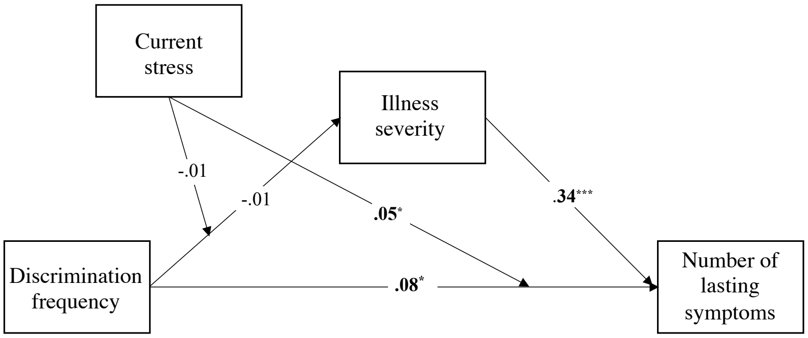


**Figure S2. Analytical control model testing moderation by general stress levels.** Current stress similarly moderates the impact of discrimination frequency on lasting symptoms, but does not moderate the direct effect of discrimination frequency on illness severity, nor the indirect association between discrimination frequency, illness severity, and lasting symptoms. All coefficients are standardized. On all pathways, we controlled for discrimination stress, non-White race, objective SES score, perceived SES score, history of mood/anxiety disorder, history of diabetes/heart disease, COVID-illness life disruption, COVID-illness anxiety, and early versus late illness onset (i.e., peak 1 versus peak 2). * $p < .05$, *** $p < .001$.

### Quality of clinical care during COVID-19 illness as a protective factor

Path analyses examined whether quality of medical care during COVID-19 illness moderated conditional associations between discrimination, illness severity, and lasting symptoms, again controlling for non-White race, objective SES score, perceived SES score, history of mood/anxiety disorder, history of diabetes or heart disease, COVID-illness life disruption, COVID-illness anxiety, and early versus late illness onset. For individuals who reported that their quality of care was not excellent ($n$ = 740; **Figure S3**), there was a significant direct association between illness severity and lasting symptoms ($\beta = .38, p < .001$), and a trending association between discrimination frequency and lasting symptoms ($\beta = .069, p = .06$). Although the direct unconditional effect of discrimination frequency on illness severity was not significant (B = -.02), the direct effect conditional on discrimination stress was significant ($\beta = .08, p = .04$). Analysis of simple slopes indicated that the effect of discrimination frequency on illness severity was more positive for individuals reporting high discrimination stress (mean +1 SD: $\beta = .01$, CI [-.003, .03]), compared to individuals reporting lower discrimination stress (mean -1 SD: $\beta = -.02$, CI [-.03, -.001]). Importantly, there was also a significant indirect association between discrimination frequency, illness severity, and lasting symptoms, which was conditional on discrimination stress ($\beta = .03$, CI [.01, .06]). Specifically, there was a greater positive indirect association between discrimination frequency, illness severity, and lasting symptoms for individuals reporting higher discrimination stress (mean +1 SD: $\beta = .03$, CI [-.01, .06]) than for individuals reporting lower discrimination stress (mean -1 SD: $\beta = -.04$, CI [-.08, -.002]). When examining individuals with excellent reported care quality ($n$ = 727; **Figure S3**), there was a significant direct effect of illness severity on lasting symptoms ($\beta = .30, p < .001$). Notably, and in contrast to individuals with non-excellent care, there were no other significant direct or indirect associations (all *βs* < .07, all *ps* > .13). These findings suggest that high quality of care may buffer associations between discrimination, illness severity, and lasting symptoms.

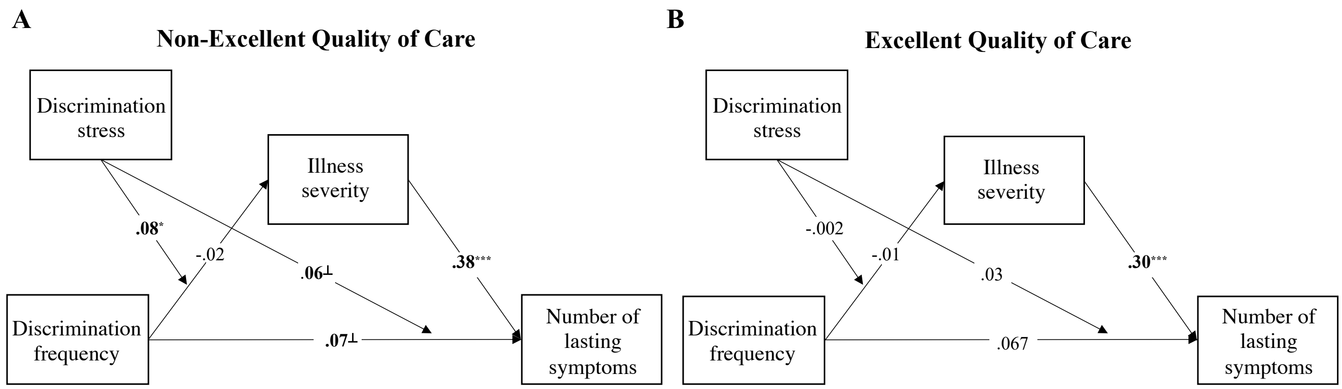


**Figure S3. In individuals reporting non-excellent care (A), there were significant direct effects of discrimination frequency on lasting symptoms, and on illness severity, which was conditional on discrimination stress.** There was also a significant conditional indirect association between discrimination frequency, illness severity, and lasting symptoms. In individuals reporting excellent care (B), there was only a direct effect of illness severity on lasting symptoms, suggesting that high quality of care may buffer associations between discrimination, illness severity, and prolonged symptoms. All coefficients are standardized. On all pathways, we controlled for discrimination stress, BIPOC race, cumulative SES risk score, perceived SES score, history of mood/anxiety disorder, history of diabetes/heart disease, COVID-illness life disruption, COVID-illness anxiety, and early versus late illness onset (i.e., peak 1 versus peak 2). ⊥ $p < .10$, * $p < .05$, *** $p < .001$.

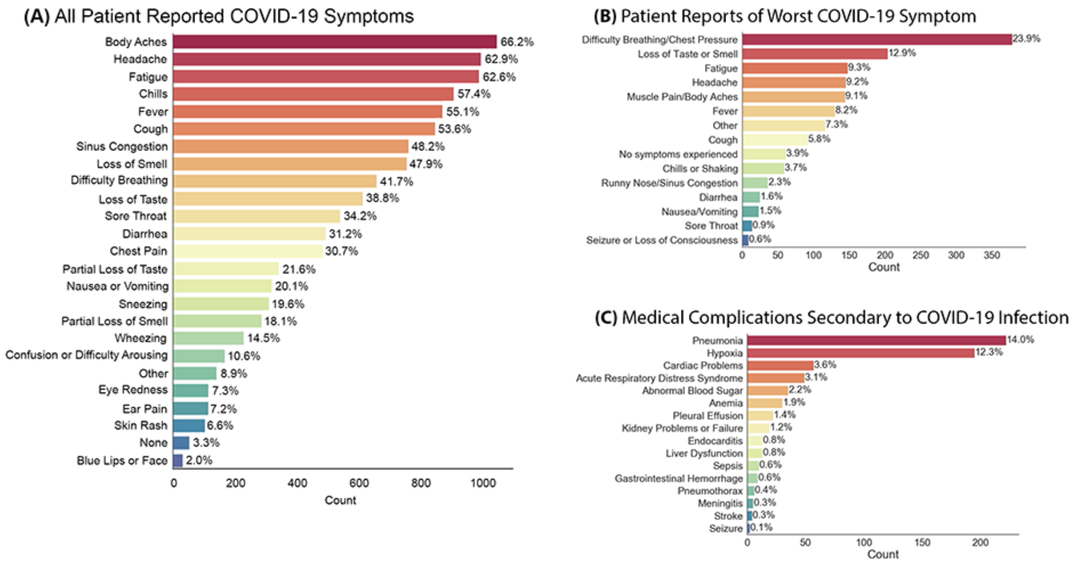


**Figure S4. Patient report of COVID-19 symptoms and related conditions.** The most prevalent concerns were chest pressure/shortness of breath, loss of taste or smell and fatigue. Pneumonia was the most frequent secondary complication of infection.

**Frequency of symptoms experienced during COVID-19 illness**

Patients were asked "What symptoms did you experience while you were ill with COVID? *(check all)*," "What was the most concerning COVID symptom or medical complication that you experienced? *(check all)*", and "What was the most concerning COVID symptom or medical complication that you experienced? *(check one)*". The most frequently endorsed concerning symptoms were difficulty breathing and loss of taste or smell.

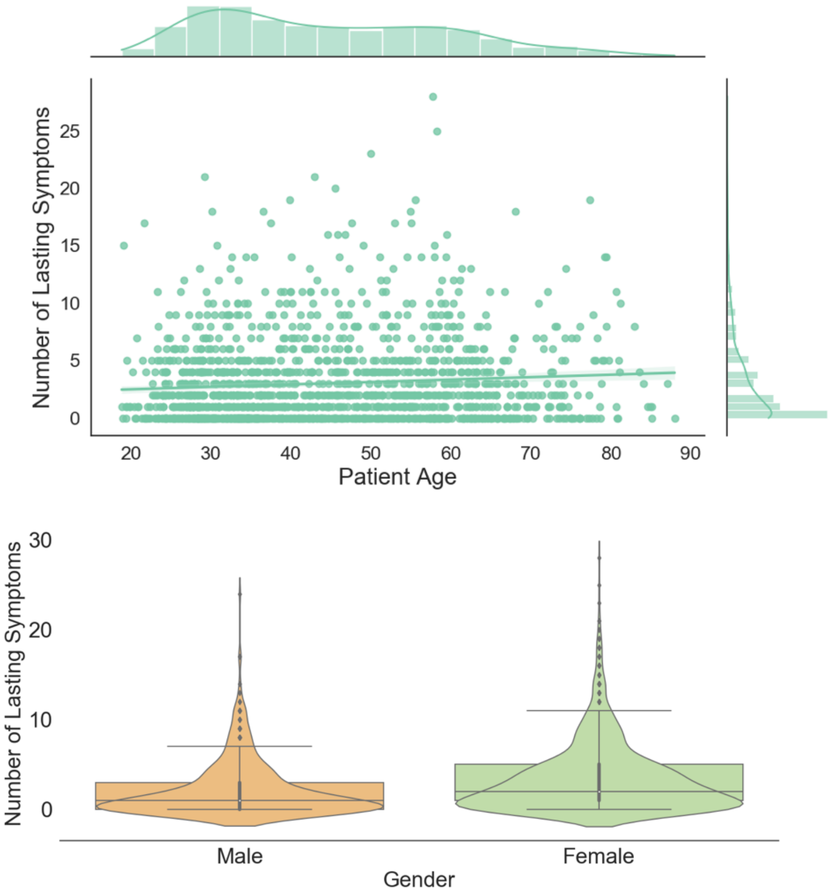


**Figure S5. Age and sex of patient relate to number of lasting symptoms after recovery from COVID-19 illness.** There was a significant association between patient age and number of lasting symptoms ($\beta$=0.07, *p*=0.01, $\Delta R^2$<0.01, *95%CI*[0.004, 0.03]) after controlling for the covariates included in our primary models: early versus late COVID-19 infection, non-White race, discrimination stressfulness, illness anxiety, illness disruption to daily life, objective SES score, perceived SES, history of mood/anxiety disorder, history of diabetes, and history of hypertension or heart disease. In addition, females reported more lasting symptoms (*M*=3.41, *SD*=3.94) compared to males (*M*=2.22, *SD*=3.02) even after controlling for the aforementioned covariates ($F$(1, 1366)=30.79, *p*=3.5E-8, $\eta^2$=0.02, *95%CI*[-1.49, -0.71]). All models were run with 5,000 bootstrapped samples.

**References**

1. Thomason, M., D. Werchan, and C. Hendrix, *COVID-19 patient accounts of illness severity, treatments and lasting symptoms.* Scientific Data, 2021.
2. Williams, D.R., et al., *Racial Differences in Physical and Mental Health: Socio-economic Status, Stress and Discrimination.* J Health Psychol, 1997. **2**(3): p. 335-51.
